# Acute rejection timing in the first post-transplant year is not associated with incident cardiac allograft vasculopathy

**DOI:** 10.64898/2026.05.28.26354171

**Authors:** Blake Butler, Shi Huang, Aniket S. Rali, Hasan K. Siddiqi, Jonathan N. Menachem, Nelson Chow, Eric Farber-Eger, Quinn S. Wells, Kelly H. Schlendorf, Kaushik Amancherla

**Author notes:** <u>**Corresponding Author:**</u> Kaushik Amancherla, MD, MSCI, Vanderbilt Translational and Clinical Cardiovascular Research Center, 2525 West End Ave., Suite 370, Office # 375, Nashville, TN 37203. **AUTHOR CONTRIBUTIONS** Conceptualization: K.A.; B.B., S.H.; Formal analysis: S.H., B.B., K.A.; Writing and revision: B.B., A.S.R., H.K.S., J.N.M., N.C., E.F.E., Q.S.W., K.H.S., K.A.

## Abstract

Heart transplantation (HT) is the durable therapy for end-stage heart failure (HF). Despite advances in immunosuppression, cardiac allograft vasculopathy (CAV) remains a leading cause of late graft failure and mortality in the modern era. Prior studies have established donor age and immunological phenomena, such as acute cellular rejection (ACR), antibody-mediated rejection (AMR), and development of donor-specific antibodies (DSAs) as risk factors for CAV^1-5^. However, it remains unclear whether acute rejection (AR) that occurs early post-HT, when individuals experience the highest degree of immunosuppression, reflects higher baseline immune activity and confers a higher risk of future CAV compared to later AR, when immunosuppression is minimized. We therefore examined whether AR occurring during pre-specified early and intermediate intervals compared to those who did not experience AR in the first post-HT year was associated with future CAV among recipients without CAV at 1 year.

## Methods

We performed a retrospective cohort study of adult HT recipients (including multi-organ recipients) at our institution between July 1, 2013 and October 31, 2023. We included individuals who were alive and free of angiographic CAV at 1-year post-HT^3^. Those with CAV detected within the first year, death before the 1-year landmark, and those who transferred care to other centers within the first post-HT year were excluded. Acute rejection (ACR: 2R/3R; AMR: pAMR1-i, pAMR2) was classified according to contemporaneous immunopathologic criteria^2,4^. Angiographic CAV was defined using standard International Society for Heart and Lung Transplantation nomenclature^3^. Early AR was defined as biopsy-proven ACR or AMR within 6 months post-HT and as intermediate AR when occurring between 6 and 12 months. The primary outcome was incident CAV. Cox proportional hazard models were constructed, including donor age and development of DSAs within 6 months and 6-12 months post-HT as covariates. Individuals without CAV were censored at the time of last coronary angiogram. This study was approved by the institutional review board.

## Results

Among 507 HT recipients meeting inclusion criteria (**Table 1**), median age was 55 years, 348 (69%) were male, and median follow-up from the 1-year landmark was ≈3 years. During follow-up, incident CAV occurred in 147 individuals. The proportion of individuals experiencing AR during early (28%) and intermediate (4.3%) intervals was lower than those who did not experience AR within the first-year (68%). In unadjusted time-to-event analyses, AR in the early or intermediate window, compared to no AR within 1-year, was not associated with future CAV (**Figure 1**). Findings were unchanged after multivariable adjustment (**Figure 2**; adjusted hazard ratio [aHR] for early vs none by 1-year comparison: 1.20, 95% confidence interval [CI] 0.83-1.74; aHR for late vs none by 1-year comparison: 1.25, 95% CI 0.65-2.38).

**Table 1.** Cohort demographics.

| Characteristic | N = 507<sup>1</sup> |
| --- | --- |
| Recipient age (years) | 55 (45, 64) |
| Donor age (years) | 30 (23, 37) |
| Male recipients | 348 (69%) |
| Time to first biopsy (days) | 8.0 (7.0, 13.0) |
| Post-landmark follow-up (years) | 2.99 (1.61, 3.22) |
| Deceased patients | 86 (17%) |
| Cardiac allograft vasculopathy | 147 (29%) |
| Donor-specific antibodies | 181 (36%) |
| Antibody-mediated rejection | 71 (14%) |
| Acute cellular rejection | 180 (36%) |
| <sup>1</sup> Median (Q1, Q3); n (%) |  |

**Figure 1.**
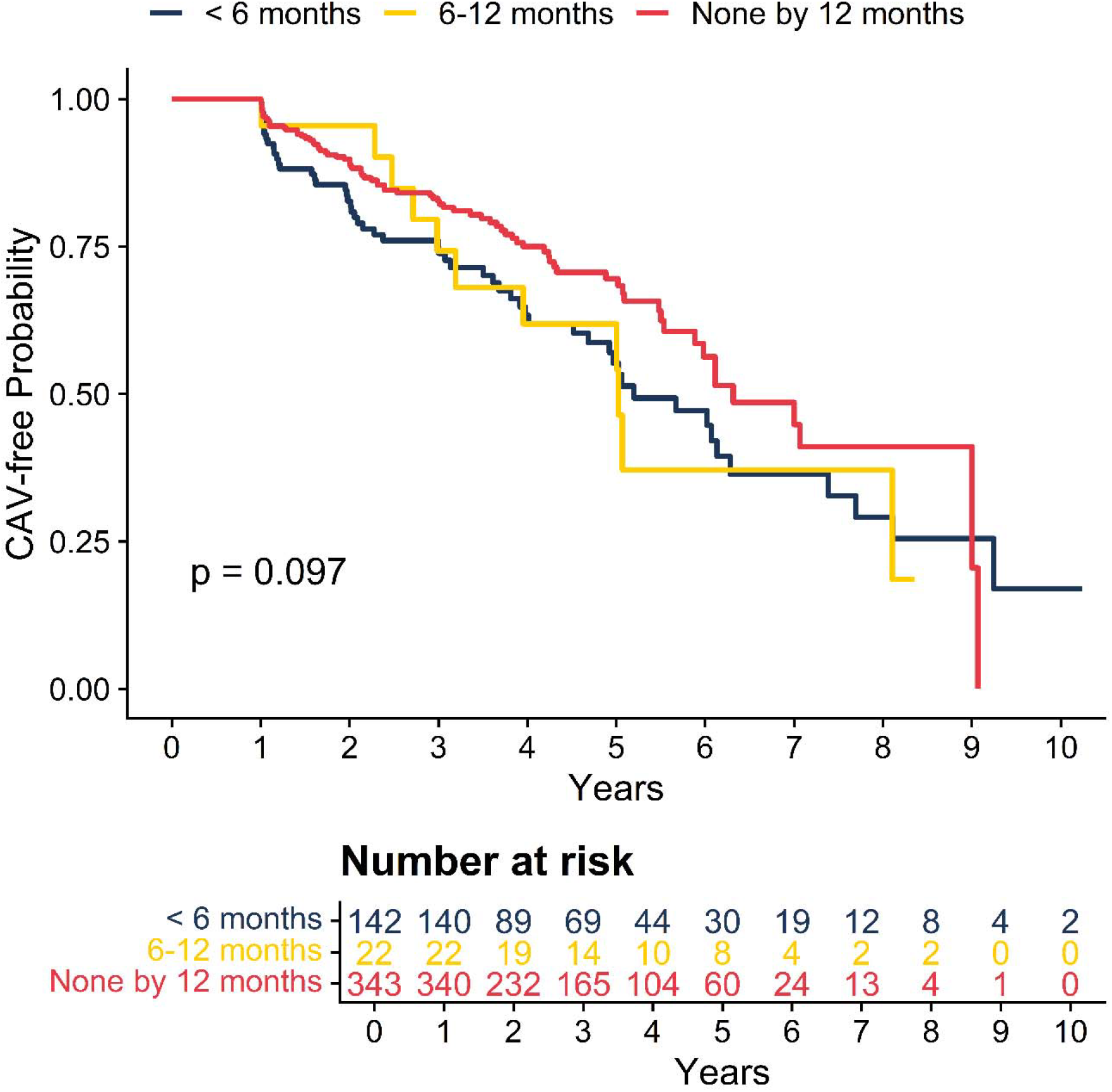
Kaplan-Meier curve of CAV-free probability over a median 2.99 year follow-up after the CAV-free 1-year landmark grouped by those who experienced at least one episode of acute rejection withing 6 months, during 6 to 12 months, and none within 12 months after transplant.

**Figure 2.**
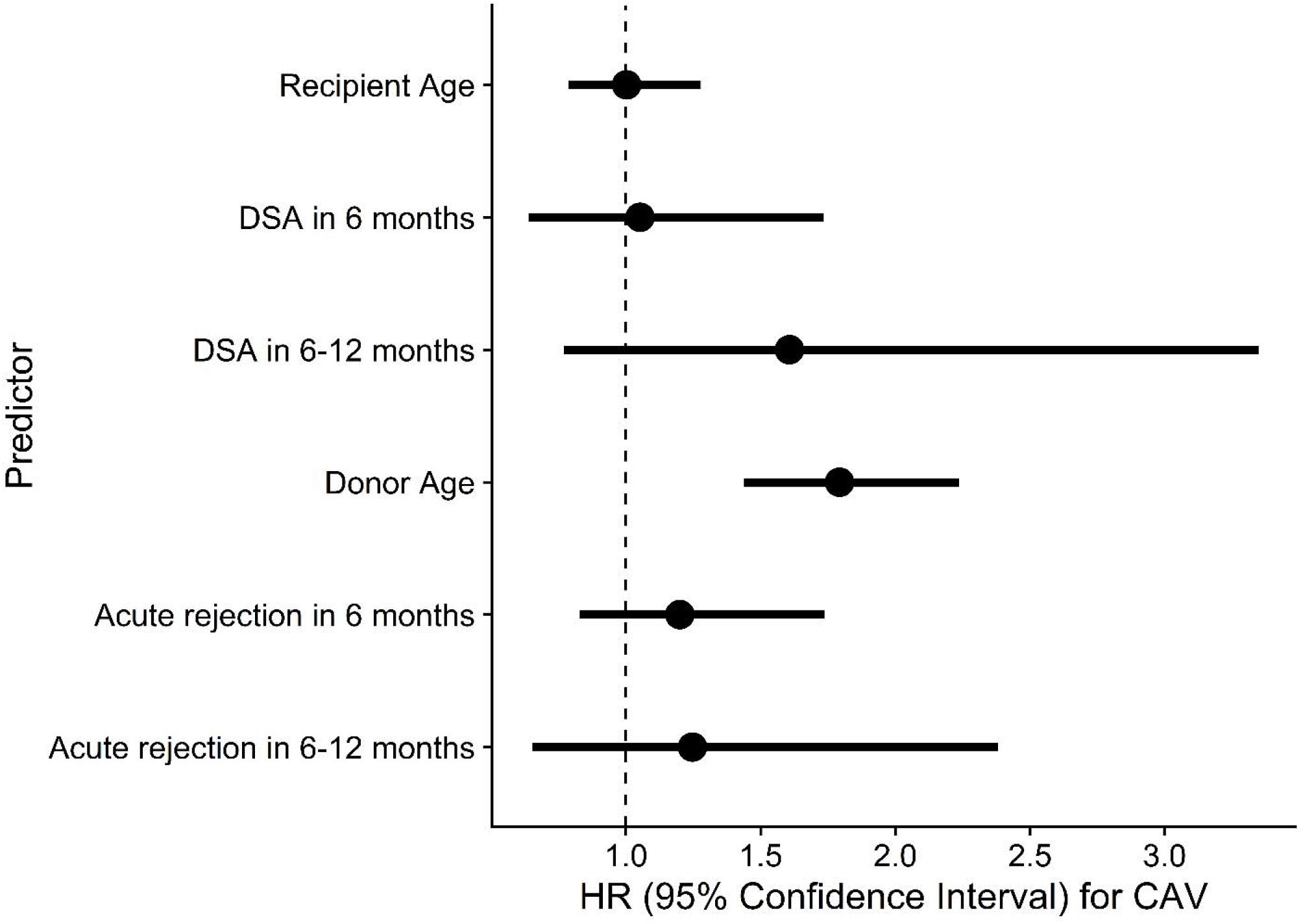
Forest plot depicted adjusted hazard ratios and 95% confidence intervals for variables tested in a Cox proportional hazards model.

## Discussion

In this landmark cohort of HT recipients free of CAV at 1 year, we did not observe an association between biopsy-defined AR during the first post-HT year and subsequent incident CAV. This finding is clinically relevant because it addresses a focused question—among recipients free of CAV at 1 year, does the timing of histologic rejection within the first year identify individuals at higher risk for subsequent CAV? In our cohort, neither early nor intermediate windows of rejection distinguished later CAV risk from those who did not experience rejection during the first post-HT year.

Our findings should also be interpreted in light of the study design. This was a single-center retrospective analysis and may have been underpowered to detect modest associations. Rejection burden was summarized within broad time windows and does not capture event frequency, treatment intensity, hemodynamic compromise, or persistent low-grade immune activity. We also used clinically available angiographic CAV surveillance rather than intravascular imaging in all patients. Nevertheless, the 1-year landmark design is a strength, as it isolates predictors of incident late CAV rather than conflating them with determinants of early vasculopathy.

In conclusion, among HT recipients free of CAV at 1 year, individuals with early rejection (within 6 months) and those with intermediate rejection (6–12 months) did not have a different risk of subsequent incident CAV compared with patients without rejection during the first post-transplant year. These data suggest that future risk stratification for CAV may require more integrated phenotyping than biopsy timing alone, including recurrent-state rejection trajectories, hemodynamics, and longitudinal biomarkers.

## Data Availability

All data produced in the present work are contained in the manuscript.

